# The effect of Alzheimer’s disease and its progression on pyramidal cell gain and connectivity

**DOI:** 10.1101/2024.04.11.24305662

**Authors:** Juliette H Lanskey, Amirhossein Jafarian, Melek Karadag, Ece Kocagoncu, Rebecca Williams, Pranay Yadav, Andrew J Quinn, Jemma Pitt, Tony Thayanandan, Stephen Lowe, Michael Perkinton, Maarten Timmers, Vanessa Raymont, Krish D Singh, Mark Woolrich, Anna C Nobre, Richard N Henson, James B Rowe, the NTAD study group

**Affiliations:** MRC Cognition and Brain Sciences Unit, University of Cambridge, Cambridge, UK; Department of Clinical Neurosciences and Cambridge University Hospitals NHS Foundation Trust, Cambridge Biomedical Campus, Cambridge, UK; Centre for Human Brain Health, School of Psychology, University of Birmingham, UK; Oxford Centre for Human Brain Activity, Wellcome Centre for Integrative Neuroimaging, Department of Psychiatry, University of Oxford, Oxford, UK; Lilly Corporate Center, Indianapolis, USA; Neuroscience, BioPharmaceuticals R&D, AstraZeneca, Cambridge, UK; Neuroscience External Innovation, Johnson & Johnson Innovations, London, UK; Department of Psychiatry, University of Oxford, Oxford, UK; Cardiff University Brain Research Imaging Centre, School of Psychology, Cardiff University, Cardiff, UK; Department of Psychiatry, University of Cambridge, Cambridge, UK

**Keywords:** dementia, canonical microcircuit, neuroimaging, effective connectivity, DCM, MMN

## Abstract

Alzheimer’s disease affects our cognitive neurophysiology by loss of neurones, synapses and neurotransmitters. An improved mechanistic understanding of the human disease will facilitate new treatments. To this end, biophysically-informed dynamic causal models can support inferences around laminar and cell-specific disease effects from human non-invasive imaging. Based on pre-clinical models and effects of cholinesterase inhibitors, we hypothesised that Alzheimer’s disease would affect the modulation of superficial pyramidal cell gain and extrinsic connectivity between pyramidal cells of different regions in hierarchical cognitive networks. Magnetoencephalography (MEG) was recorded during an auditory mismatch negativity task from healthy adults (n=14) and people with symptomatic Alzheimer’s disease or mild cognitive impairment (n=45, all amyloid-biomarker positive) at baseline and after 16 months. Fourteen people from the symptomatic group had repeat magnetoencephalography at two weeks to assess test-retest reliability. Sensor-level data were analysed using t-tests of the mismatch negativity amplitude from 140ms to 160ms. The repetition effect was assessed with repeated-measures analysis of covariance, using the average evoked response in the mismatch negativity time window as the repeated measure. An absolute, intraclass correlation model of the test-retest data assessed mismatch negativity amplitude reliability. We then fitted dynamic causal models to the evoked responses over 500ms. Second-level parametric empirical Bayes across participants examined the effect of (1) group, patients vs controls, and (2) progression, baseline vs follow-up, on the model parameters reflecting pyramidal cell gain modulation and extrinsic connectivity. There was a significant effect of both disease and progression on the mismatch negativity amplitude (patients vs controls, T=-1.80, p=0.04; patient baseline vs follow-up, T=-2.72, p=.005), which had excellent reliability (ICC=0.95, p<.001). Parametric empirical Bayes revealed strong evidence (posterior probability>95%) that Alzheimer’s disease reduced extrinsic connectivity and superficial pyramidal cell gain modulation, which was reduced further at follow up assessment. The mechanistic modelling confirmed the hypothesis that reduced superficial pyramidal cell gain modulation and extrinsic connectivity can explain the observed neurophysiological effect of Alzheimer’s disease. This approach to non-invasive magnetoencephalography data may be used for experimental medicine studies of candidate treatments, and bridge clinical to preclinical models of drug efficacy.

## Introduction

Developing effective treatments for Alzheimer’s disease remains a major challenge (Anderson *et al*., 2017). Effective treatments should target the disease mechanisms behind the neurophysiological and cognitive deficits (Yiannopoulou *et al*., 2019). To develop these treatments *in vivo,* human assays that quantify the causes of neurophysiological change are needed (Drummond and Wisniewski, 2017). Here, we test the sensitivity of the inversion between magnetoencephalography and generative models of human neurophysiology to the presence and progression of Alzheimer’s disease. This approach allows human, *in vivo* characterisation of neural dynamics by cortical layer and cell type from non-invasive neuroimaging (Friston *et al*., 2003).

We focus on the impact of Alzheimer’s disease on the connectivity of pyramidal cells. Pyramidal cells in superficial layers of cortex are modulated by currently licenced cholinergic treatments for dementia (Moran *et al*., 2013). For example, galantamine modulates activity of superficial pyramidal cells, leading to changes in neurophysiological responses to rapidly changing stimuli. In rodent models, cholinergic agonists and antagonists have opposing dose-dependent effects on superficial pyramidal cell gain modulation (Schöbi *et al*., 2021). The hippocampus, which has early and significant involvement in the course of Alzheimer’s disease, may be involved in inhibiting and disinhibiting superficial pyramidal cells through regulation of acetylcholine (Barron *et al*., 2020).

Pyramidal cells are directly affected by the molecular hallmarks of Alzheimer’s disease: extracellular beta-amyloid plaques (Braak and Braak, 1997) and intraneuronal tau tangles (Braak and Braak, 1991; Thangavel *et al*., 2008). Tau aggregates develop in pyramidal cells and can spread between pyramidal cells (Braak and Tredici, 2018). Variance in the distribution and burden of tauopathy in Alzheimer’s disease is closely associated with the clinical phenotype (Ossenkoppele et al., 2016), cognitive deficit (Nelson et al., 2012) and rate of cognitive decline (Malpetti *et al*., 2020; Tanner *et al*., 2022). The effect of aggregated tau on cognition in part results from synaptic toxicity and dendritic dearborization of pyramidal cells (Allard *et al*., 2012; Merino-Serrais *et al*., 2013; Mijalkov *et al*., 2021).

Identification of pyramidal cell function in humans, *in vivo,* can be achieved by invasive electrocorticography, usually in people undergoing evaluation of epilepsy. This is not practical as a basis for research and drug development in the context of Alzheimer’s disease. An alternative approach is the inversion between functional neuroimaging (e.g., electro- and magneto-encephalography, E/MEG) and biophysical models of the cortex. This approach has been used to study the effects of anti-NMDA autoimmune encephalopathy (Symmonds et al., 2018), inherited channelopathies (Gilbert et al., 2016), ageing (Moran et al., 2014), pathophysiology of non-Alzheimer dementias (Shaw *et al*., 2019; Adams *et al*., 2020, 2021, 2023) and confirmatory studies of drug interventions (Moran *et al*., 2013; Adams *et al*., 2021).

Here we use generative models with non-invasive human MEG to test the hypothesis that Alzheimer’s disease affects pyramidal cell gain and connectivity, which influence the neurophysiological response to unexpected inputs. We do so in the context of a sensory mismatch negativity paradigm, in which participants automatically learn and adapt to changing auditory stimulus frequency. This learning reflects short-term plasticity (Garrido *et al*., 2009) and is achieved by iterative updating of top-down predictions by bottom-up prediction errors (Auksztulewicz and Friston, 2016). As top-down predictions of a repeated stimulus become more accurate with short-term learning, prediction errors (and their neurophysiological correlate) reduce. With repetition, the network becomes more precise in its predictions such that prediction errors acquire greater precision (Auksztulewicz and Friston, 2016). An initial deviant stimulus retains the high precision of the previous standard, thereby enhancing the signal of the large prediction error (i.e., a large magnitude error made with high precision). Following such a large prediction error, the precision (c.f., the confidence, in lay terms) in the model is reduced, such that the subsequent prediction error is less precise and generates an attenuated mismatch response.

The precision of prediction errors is proposed to be encoded by the gain modulation of superficial pyramidal cells (Moran *et al*., 2013; Schöbi *et al*., 2021), while prediction error is proposed to be mediated by superficial pyramidal cell forward connections (Auksztulewicz and Friston, 2016). In Alzheimer’s disease, this mechanism is disrupted with changes including reductions in precision and prediction error signalling (Kocagoncu *et al*., 2021). Indeed, novelty encoding (Bastin *et al*., 2019) and auditory predictions (Pérez-González *et al*., 2022) are disrupted in Alzheimer’s disease, particularly in parietal and medial temporal cortices (Billette *et al*., 2022; Pérez-González *et al*., 2022). This deficit would not only impair learning, but also signal processing and perception in a noisy environment, as observed in Alzheimer’s disease (Pérez-González *et al*., 2022).

Our overarching hypothesis is that the observed physiological effect of Alzheimer’s disease on the mismatch negativity response is explained by reduced gain modulation of superficial pyramidal cells and reduced effectiveness of extrinsic connectivity between pyramidal cell populations of connected regions. We test two specific predictions: (1) symptomatic Alzheimer’s disease, in the form of amyloid-positive mild cognitive impairment and mild Alzheimer-type dementia, is associated with reduced gain modulation of superficial pyramidal cells and reduced extrinsic connectivity between pyramidal cells; (2) the same features of the cortical microcircuit change with disease progression.

## Materials and methods

### Participants

Data were drawn from the New Therapeutics in Alzheimer’s Disease (NTAD) multicentre longitudinal observational study (Lanskey *et al*., 2022). The study included neurologically-normal, control participants and participants with a clinical diagnosis of Alzheimer’s disease or mild cognitive impairment. Participants completed (i) amyloid screening through either positron emission tomography or cerebrospinal fluid collection, a positive amyloid status enabled study participation for prospective patient participants; (ii) a battery of clinical and neuropsychological assessments; and (iii) neuroimaging, comprising MRI and E/MEG protocols. People in the control group completed screening and baseline assessments only, while people in the patient group completed additional, longitudinal assessments as follows: clinical and neuropsychological assessments were completed at three timepoints with 12-16 months between sessions; blood collection, E/MEG and MRI were completed at baseline and again at 12-16 months. An additional E/MEG scan was completed two-weeks after baseline for a subset of people from the patient group (see Lanskey *et al*., 2022 for further details).

For the current analysis, we used the MEG, MRI and demographic baseline data from participants who completed the mismatch negativity MEG recording. Of the 50 people with mild cognitive impairment or Alzheimer’s disease who completed the NTAD baseline MEG scan at the Cambridge site, we excluded two people who did not complete the mismatch negativity task as the earphones did not fit comfortably, two people whose diagnosis was revised during follow-up and one due to data recording technical issues at baseline. Of the 15 people with mild cognitive impairment or Alzheimer’s disease who completed the test-retest MEG scans at the Cambridge site, one person was excluded from the analysis because of trigger-failures on the retest session. One control participant was amyloid-positive and excluded from the current analysis. The resulting participant numbers comprised 45 people with Alzheimer’s disease or mild cognitive impairment with a positive amyloid status and 14 neurologically-normal control participants. For the test-retest analysis, we used the subset of 14 participants from the test-retest subgroup of participants with Alzheimer’s disease or mild cognitive impairment. Clinical and cognitive data for participants are given in Table *1*. Independent Student’s t-tests and Bayesian t-tests were used for continuous data, or chi-squared tests for categorical data, to identify differences at baseline between participants with Alzheimer’s disease and control participants.

**Table 1.**
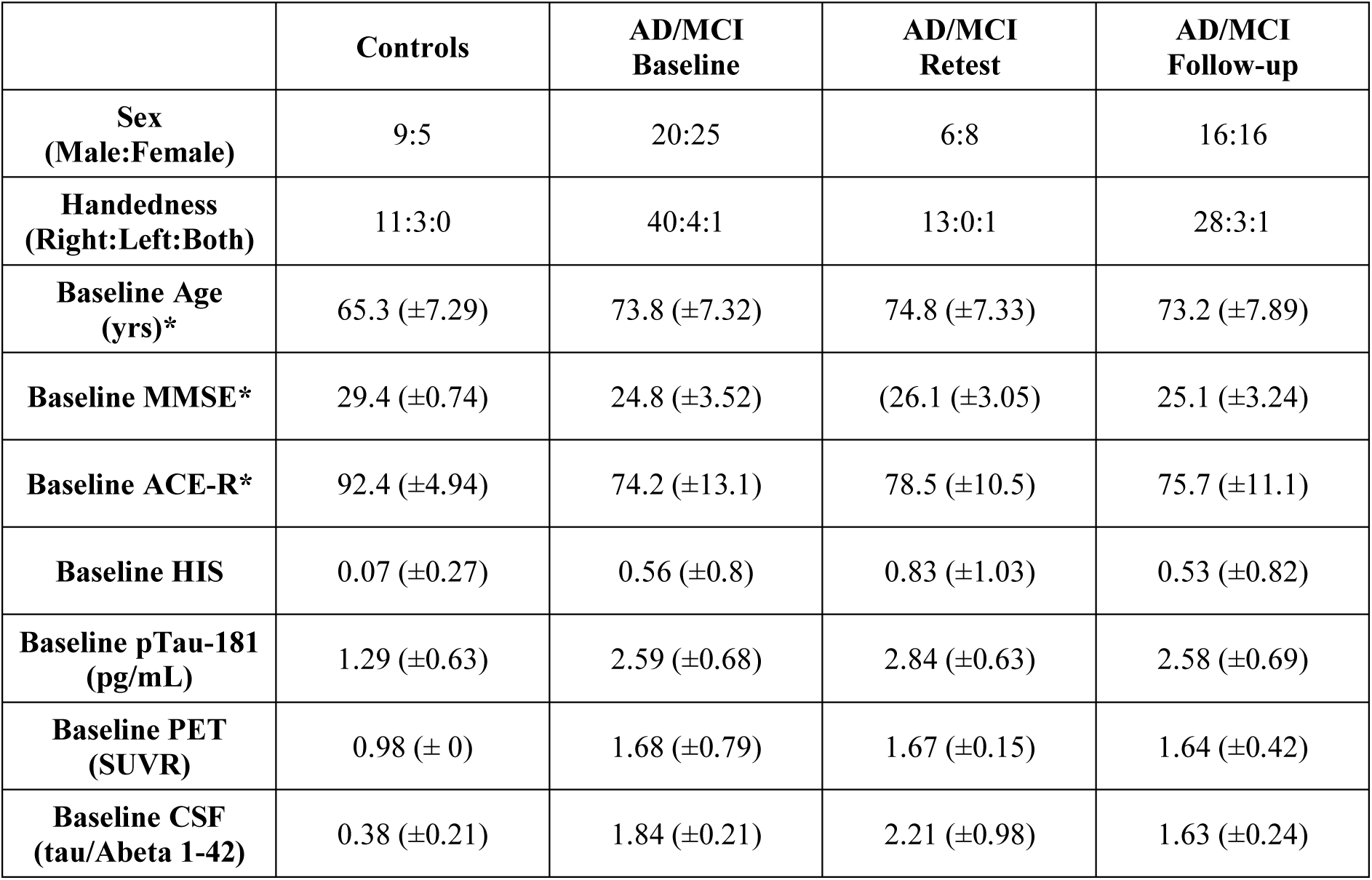
Participant demographics and clinical data. Values are given as mean (standard deviation) *denotes significant difference between baseline groups by frequentist and/or Bayesian statistics, as per text. Yrs, years; MMSE, mini-mental state examination; ACE-R, Addenbrooke’s cognitive examination revised; HIS, Hachinski ischaemic score; PET, positron emission tomography; SUVR, standardised uptake value ratio; CSF, cerebrospinal fluid

### Task

We used a passive roving mismatch negativity task (Adams *et al*., 2020) while the participants passively watched a muted movie. The auditory mismatch negativity task elicits responses to unexpected ‘oddball’ stimuli followed by rapid plasticity as predictions are updated upon repetition of the new stimulus (Friston, 2005; Kiebel et al., 2007). Through non-magnetic earpieces, the participants hear blocks of short sinusoidal tones binaurally in phase, 60dB above the average auditory threshold. The tones are presented for 100ms at 500ms intervals with frequencies in the range 400 to 800Hz varying in 50Hz steps. The frequency of successive tones is the same within blocks but changes between blocks. The number of repeated tones per block varied from 3 to 11. Consequently, the first tone of each block represents a deviant tone, which becomes a new standard tone upon repetition.

### Imaging acquisition

MEG data were collected on the ElektaVectorView system and MEGIN Triux Neo scanner. Both scanners were configured with 204 planar gradiometers and 102 magnetometers. The position of five head position indicator coils, standard fiducial points and over 300 additional head points were recorded using the Polhemus digitization system. ECG data was recorded by an electrode on the right clavicle and a second electrode on the left, lower rib. A reference electrode recorded from the left side of the nose and the ground electrode was placed on the left cheek. Electrodes on bilateral canthi and below and above the left eye recorded electro-oculogram data.

T1-weighted MRI was recorded from each participant with a 3T Siemens PRISMA scanner using a magnetisation prepared rapid gradient echo (MPRAGE) sequence (echo time, 2.91ms; inversion time, 900ms; repetition time, 2300ms; flip angle, 9 degrees; voxel size, 1mm isotropic; slice thickness, 1mm; slice number, 176; acquisition matrix, 256×240).

### Data analyses

#### Pre-processing

MaxFilter v2.2 software (Elekta Neuromag) was used on the raw data to automatically detect bad channels, perform temporal signal space separation, and correct for head movement. Independent component analysis was performed using the EEGLAB toolbox (Delorme and Makeig, 2004) to detect and remove artefactual components that correlated with EOG and ECG timeseries using cardiac spatial and normative blink templates. The residual timeseries were subsequently reconstructed for further preprocessing using SPM12 (r7771). Data were bandpass filtered between 0.01Hz and 40Hz and epoched from −100ms to 500ms relative to stimulus onset (0ms). OSL’s artefact rejection algorithm (github.com/OHBA-analysis/osl-core) was used to remove residual bad channels and trials. Robust averaging was used to average epochs for deviant and repeated trials, with conditions weighted separately. A final low-pass filter was applied to the data to correct for potential high frequencies introduced during robust averaging.

#### Sensor space analysis

For the sensor space analysis, planar gradiometers were combined by calculating their root mean square. We calculated the mean response across planar gradiometers for each subject, for each trial type, for each session separately (baseline, two-weeks and annual follow up). The repetition effect was assessed with a repeated-measures ANCOVA, using the average evoked response in the mismatch negativity time window (140-160ms, Moran *et al*., 2013) with tone repetition number (deviant, repetition 1, repetition 2, repetition 3, repetition 4 and repetition 5) as the repeated measure, group (controls versus patients) for the between-subject measure and age as the covariate. We also assessed the difference waveform for each subject; the response to the deviant tone was subtracted from the response to the first stimulus repetition. We calculated the average amplitude of this difference waveform from 140 to 160ms. An independent sample t-test was used to compare means of the mismatch negativity amplitude between groups. Group average waveforms were plotted after averaging difference waveforms across participants of each group.

For the longitudinal patient data, the difference in means between baseline and follow-up sessions was assessed using a t-test. The repetition effect was assessed with a repeated-measures ANOVA, using the average evoked response in the mismatch negativity time window (140-160ms, Moran *et al*., 2013) with tone repetition number (deviant, repetition 1, repetition 2, repetition 3, repetition 4 and repetition 5) and session (baseline versus follow-up) as repeated measures.

We estimated test-retest reliability of the baseline and two-week data using an absolute, intra-class correlation model (Koo and Li, 2016). With similar means, the intra-class correlation coefficient is numerically equivalent to the product moment correlation (estimated as Pearson’s correlation coefficient).

#### First level network modelling

We used dynamic causal modelling with laminar-specific parameters to determine the effect of Alzheimer’s disease on the neurophysiological generators. We used a canonical microcircuit model for evoked responses (Bastos *et al*., 2012; Pinotsis *et al*., 2013) comprising of four cell populations at each of 8 cortical regions (see **Figure 2**): bilateral primary auditory cortex (left MNI coordinates: −42, −22, 7; right MNI coordinates: 46, −14, 8), bilaterial superior temporal gyrus (left: −61, −32, +8; right: +59, −25, +8), bilateral inferior frontal gyrus (left: −46, +20, +8; right: +46, +20, +8), and bilateral inferior parietal cortex (left: −58, −27, +30; right: +59, −41, +30). Six of these regions are based on a series of prior studies with healthy adults and those with non-Alzheimer dementia (Garrido *et al*., 2008; Phillips *et al*., 2015; Rosch *et al*., 2019; Adams *et al*., 2021). We extended the network to include the inferior parietal cortex (Lappe *et al*., 2013), given the impact of Alzheimer’s disease on parietal cortex (Jacobs *et al*., 2012; Talwar *et al*., 2021) and recent evidence of mismatch negativity responses from parietal cortex (Lappe *et al*., 2013; Cope *et al*., 2022). We defined multiple, alternative, symmetric model architectures, each representing plausible alternative hierarchical relationships between regions and their connectivity to include the inferior parietal cortex (**Figure 2**). Given that higher regions send backward connections to, and receive forward connections from, lower regions in a hierarchical network, we specified forward and backward connections according to the following hierarchy as set out previously (Garrido *et al*., 2008; Phillips *et al*., 2015; Rosch *et al*., 2019; Adams *et al*., 2021): inferiFor frontal gyrus > superior temporal gyrus > primary auditory cortex. These connections were unchanging across models. All models included auditory input to primary auditory cortex and expectancy inputs to inferior frontal cortex (Phillips *et al*., 2015). Models 1 and 4 had no connections between frontal and parietal nodes. Models 2 and 5 had the inferior parietal cortex higher than the inferior frontal cortex in the cortical hierarchy. Models 3 and 6 had the inferior frontal cortex higher than parietal cortex in the cortical hierarchy. Models 1-3 had no expectancy input to parietal cortex, while models 4-6 had expectancy input to parietal cortex **Figure 2B**. The laminar asymmetry of forward vs backward connectivity is based on the canonical microcircuit model described by Bastos and colleagues (2012).

We modelled the first 6 repetitions of tones in the mismatch negativity paradigm to identify the effect of repetition on parameters from the first tone of each block (deviant) to the 6^th^ tone (a new ‘standard’ for each block). In anticipation of short-term plasticity within the dynamic causal models, we modelled the changes to neural responses over successive tone repetitions as varying by (1) exponential decay or (2) exponential decay combined with phasic change (as used by Garrido and colleagues, 2009). The second set of models with both exponential and phasic change embody the hypothesis of predictive coding for sequential neural responses to stimuli, allowing extrinsic connections to show stimulus-specific adaptation with exponential decrease with repetition; while intrinsic connections show a phasic response, reducing after the deviant tone and recovering with subsequent tones thereby reflecting the precision of prediction error (Garrido *et al*., 2009; Rosch *et al*., 2019).

The neuronal parameters for each participant’s dynamic causal model were inferred from their observed response fields of gradiometers using a lead field informed by subject-specific T1 magnetic resonance images. This inversion uses the Bayesian variational Laplace method that minimizes the free energy of the model as a cost function to estimate the free energy of the model’s and posterior estimates of its parameters (Friston *et al*., 2003, 2007). Of the 14 controls and 45 patient participants included at baseline, one control and five patients had an inversion failure during model fitting and were excluded from the parametric empirical Bayes (PEB) analyses. For the longitudinal session, three patients failed to invert during model fitting and were excluded from the longitudinal parametric empirical Bayes analyses.

Bayesian model reduction was applied to each individual’s dynamic causal model to identify the most likely explanatory model for group differences (the “winning” model) from the model space. The model space included 12 canonical microcircuit models (**Figure 2A**), with 2 sets of the 6 alternative extrinsic connectivity architectures among eight regions; one set describing the effect over sequential tones with an exponential decay and the second set, as a combination of exponential and phasic basis functions (Garrido *et al*., 2009). The free energies of reduced models were averaged (i) across all participants at baseline and (ii) separately, across all baseline and longitudinal data for patient participants. The same model had the highest model evidence for both baseline and longitudinal datasets. This model with the highest model evidence was taken forward to PEB analysis to study group differences.

#### The effect of Alzheimer’s disease

Second-level analysis with PEB of the winning model examined how disease group (healthy control *vs* Alzheimer’s disease) affected the change in generative model of successive repetitions after a change in tone frequency. We ran two analyses with PEB. For both, the PEB design matrix comprised a unitary regressor for the group mean (ones) and a regressor for group membership (zeros for control participants and ones for people with Alzheimer’s disease or mild cognitive impairment).

The first PEB analysis tested whether individual differences in the gain of superficial pyramidal cells explains the group difference in the scalp response to the mismatch negativity task. The gain modulation parameter represents a lump sum of features such as potassium conductances and hyperpolarisation after an action potential, which act as an auto-regulation or self-inhibition of the cell population (rather than being a biological inhibitory connection per se; Moran *et al*., 2013). A second PEB analysis tested the effect of individual differences in the extrinsic connectivity between pyramidal cells between all regions (both forward and backward connections). For both PEB models, Bayesian model comparison and averaging was performed over a model space of all parameter combinations with hemispheric symmetry and the Bayesian model average results were plotted.

#### The effect of Alzheimer’s disease progression

For the analysis of longitudinal change, we performed two second-level analyses with PEB applied to baseline and annual follow-up data from patients. Both PEB models included a group mean regressor and a regressor specifying whether the scan was at baseline, given a value of 0, or at the annual follow-up, given a value of 1. A third regressor specified the time in years between the baseline and follow-up scans for each patient (0 for baseline scans and with scores from 0.8 to 2.2 for follow-up scans mean-centred and standardised). Two such PEB models were inverted to examine the impact of disease progression on the (i) gain modulation of superficial pyramidal cells and (ii) extrinsic connectivity between superficial and deep pyramidal cells. For each PEB model, we performed Bayesian model comparison and averaging over a reduced model space of only those parameters that had differentiated patients *versus* controls at baseline, and then the Bayesian model average results were plotted.

#### Data availability

The code used is available at https://github.com/jlansk/dcm_cmc_ntad. Anonymised (unlinked) raw data will be made available via Dementias Platform UK, subject to managed access conditions that protect participant confidentiality and conditions of consent.

## Results

Demographic and clinical data are shown in Table 1. There were no significant differences between the groups in terms of sex (χ²=0.98, p=.32), handedness (t=1.86, p=.40), or Haschinski ischaemic score (χ²=5.12, p=.16). Patients were on average older than controls (t=-3.80, p=<.001, BF=72). As expected, there were group differences for the mini-mental state examination (t=4.75, p<.001, BF=1163), ACE-R (t=5.02, p<.001, BF=2722), and clinical dementia rating (χ²=55, p<.001). The baseline data of those completing two-week retest magnetoencephalography, and annual follow-up, were similar to the whole baseline group.

### Sensor-level effects of Alzheimer’s disease

The amplitude of the mismatch response from 140 to 160ms was significantly reduced in patients compared to controls at baseline (T = −1.80, p = 0.04, ***Figure 1B*** upper panel). The repeated-measures ANCOVA found a significant main effect of repetition (F(5, 280) = 38.0, p<0.001); that interacted with group (F(5, 280) = 2.48, p = 0.03), where the responses were smaller for patients particularly for early repetitions.

**Figure 1.**
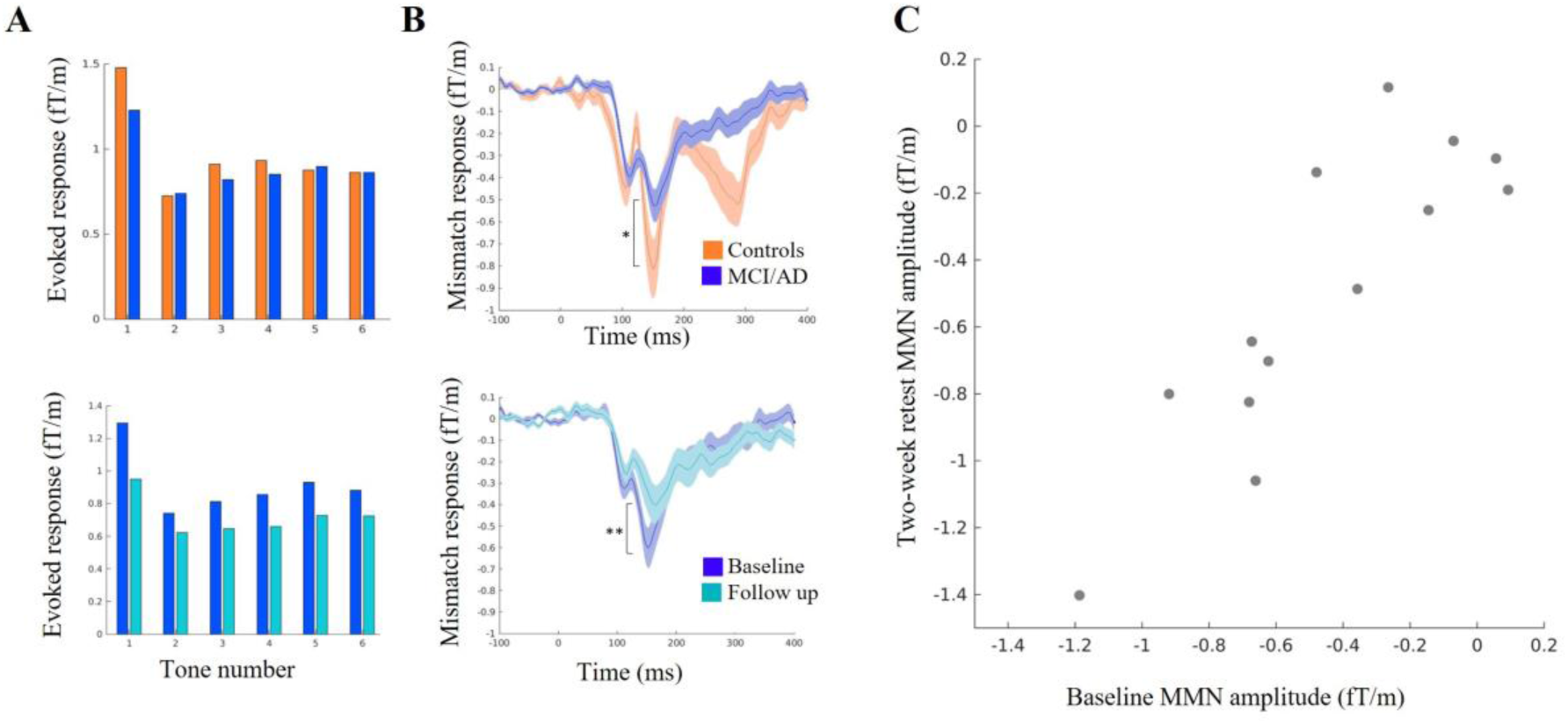
Scalp responses to the mismatch negativity task. **(A)** Amplitude of the evoked scalp responses to tones 1-6, averaged across root mean square values of all gradiometers and across samples within the *a priori* mismatch negativity window from 140ms to 160ms; top panel - for baseline controls in blue and patients in orange; bottom panel - for patients at baseline in dark blue and follow-up in light blue. **(B)** Average mismatch negativity waveform of repetition 1 minus deviant tones; top panel - for baseline controls in blue and patients in orange; bottom panel - for patients at baseline in dark blue and follow-up in light blue. (**C**) Absolute intraclass correlation for the mismatch negativity calculated using the test-retest data. MCI/AD, mild cognitive impairment or Alzheimer’s disease; MMN, mismatch negativity

The patients’ mismatch response was further attenuated (less negative) at annual follow up compared to patients’ baseline (T = −2.72, p = .005, ***Figure 1B*** lower panel). In the longitudinal repeated-measures ANOVA, there was a significant main effect of tone repetition (F(5, 155) = 20.0, p<.001) and progression (baseline versus follow-up, F(1, 31) = 75.2, p < .001), which interacted with each other (F(5, 155)=16.6, p<.001). The mismatch negativity amplitude had high reliability across the baseline and two-week test-retest sessions (ICC = 0.95, p < .001, ***Figure 1*C**).

### Dynamic causal models

The evoked responses generated by dynamic causal models were accurate, with an average Pearson correlation between generated and observed responses of 0.88 (±1.70) and 0.84 (±0.15) for patients at baseline and follow-up, respectively; and 0.85 (±0.18) for controls at baseline (see the supplementary figures for individual model fits).

The model with the highest model evidence was the same for (1) models across baseline patient and control groups (see **Figure 2C**) and (2) models across baseline and follow-up scans for patients (see **Figure 2D**). The winning model identified the inferior frontal cortex as above the parietal cortex in the network hierarchy. The winning model had the combination of exponential and phasic basis functions changes in parameters over successive repetitions, in accordance with (Garrido *et al*., 2009; Rosch *et al*., 2019) and in line with a predictive coding framework for the mismatch negativity task rather than stimulus-specific adaptation only (Garrido *et al*., 2009).

**Figure 2.**
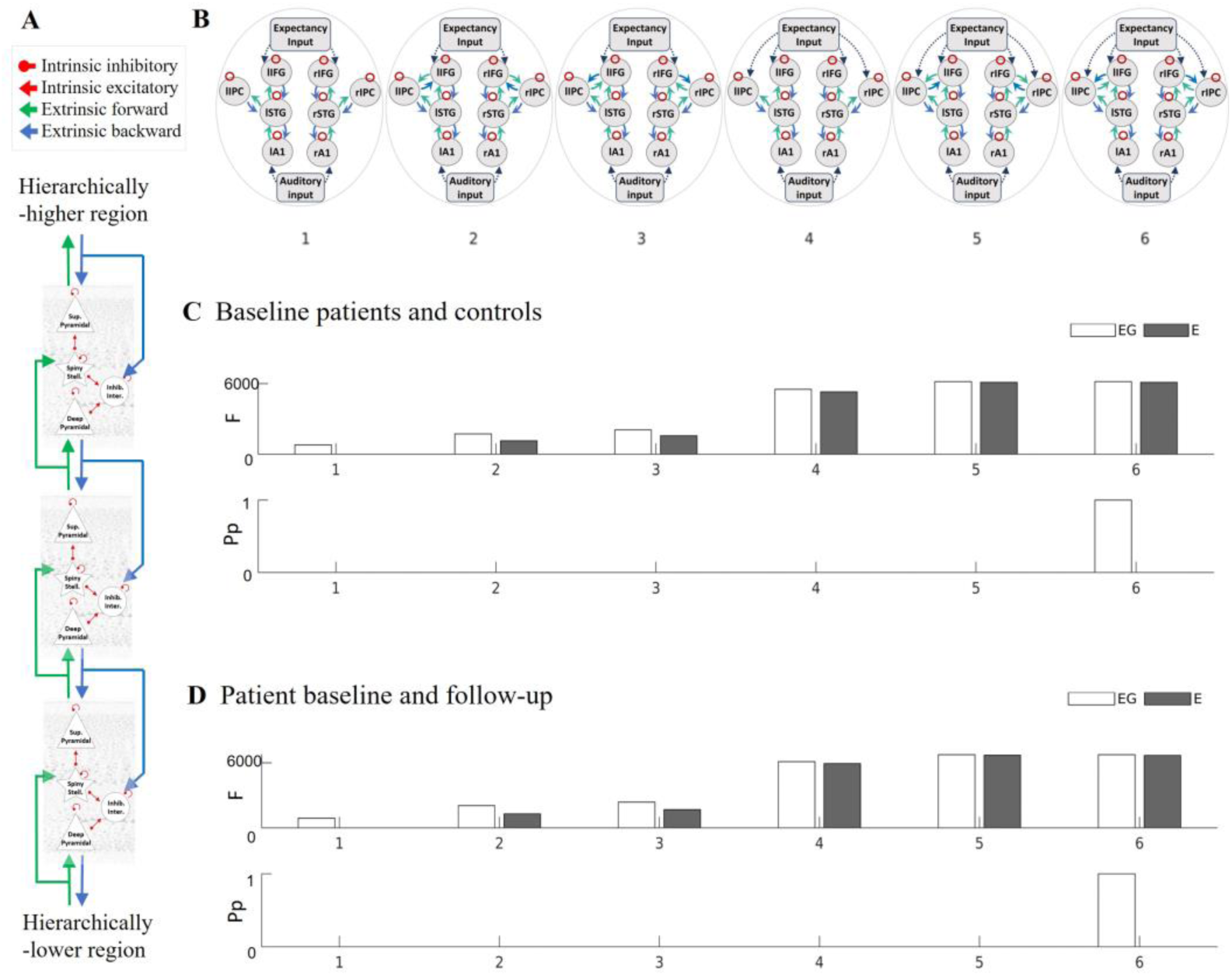
Defining the model structure. **(A)** The canonical microcircuit model for evoked responses; **(B)** Model space for determining the connectivity of the inferior parietal nodes; **(C)** The free energy (top panel) and posterior probability (bottom panel) of models for the patient and control groups; **(D)** and for the patient baseline and follow-up groups. Sup, superficial; stell., stellate; inter., interneurons; inhib., inhibitory; F, free energy; Pp, posterior probability; l, left; r, right; A1, primary auditory cortex; STG, superior temporal gyrus; IFG, inferior frontal gyrus; IPC, inferior parietal cortex

### Superficial pyramidal cell gain modulation is reduced by Alzheimer’s disease

The analyses across all regions confirmed that Alzheimer’s disease (**Figure 3B**: control versus patient PEB model) reduced the gain modulation of superficial pyramidal cells in auditory cortices and inferior frontal gyri. Disease progression also negatively modulated the gain of superficial pyramidal cell self-inhibition in the inferior frontal cortex (**Figure 3*C***: patient baseline versus follow-up PEB model).

**Figure 3.**
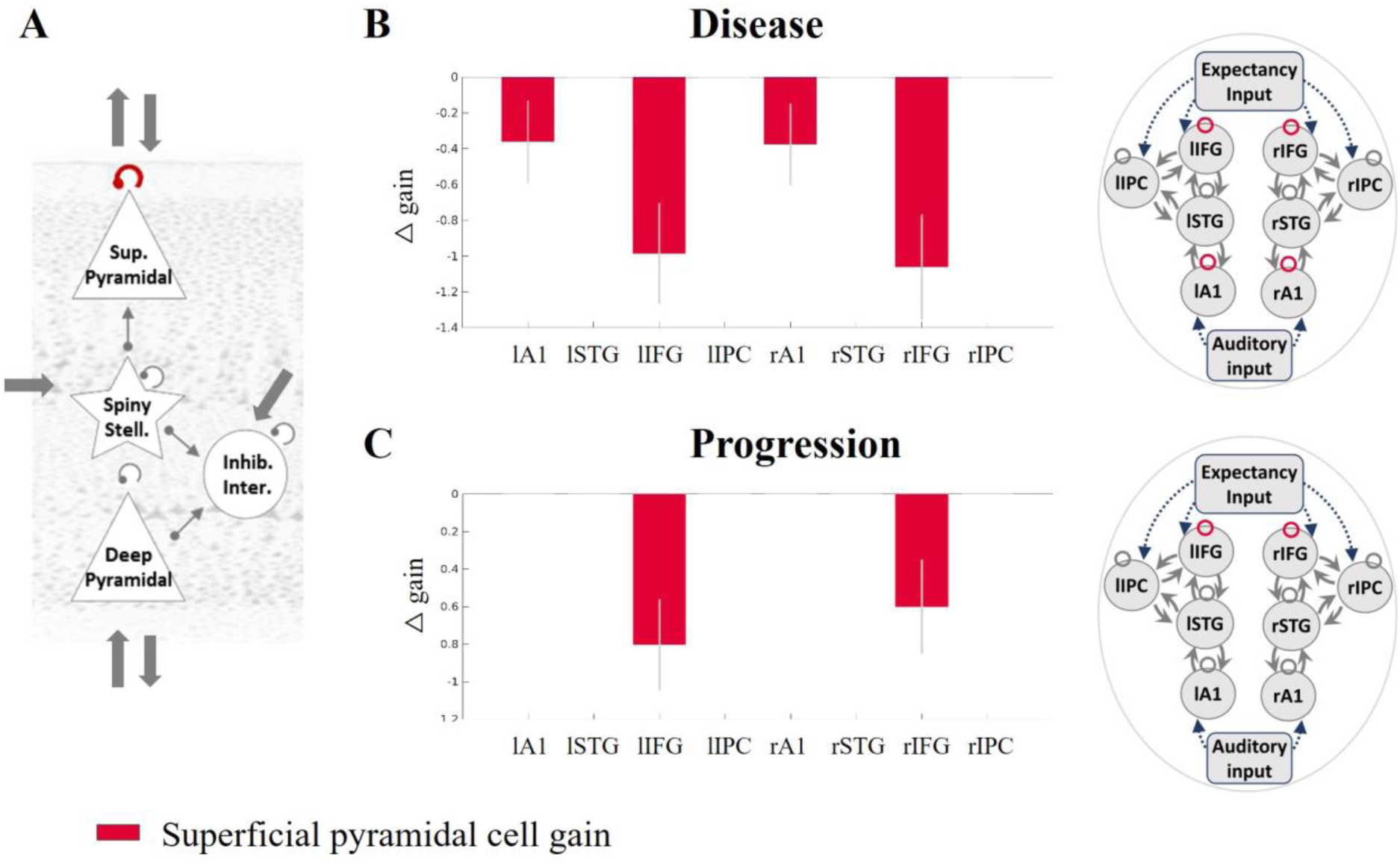
Gain of the superficial pyramidal cell self-inhibition and the effect of disease and progression. **(A)** The superficial pyramidal cell self-inhibitory connection is shown in red. Regions where this connection was modulated with a posterior probability >0.95 by **(B)** disease presence and **(C)** progression are shown in the in red. Sup, superficial; stell., stellate; inter., interneurons; inhib., inhibitory; F, free energy; Pp, posterior probability; l, left; r, right; A1, primary auditory cortex; STG, superior temporal gyrus; IFG, inferior frontal gyrus; IPC, inferior parietal cortex

### Extrinsic connectivity: effect of disease and progression

The analyses assessing extrinsic connectivity between superficial and deep pyramidal cells confirmed that the effect of disease was to weaken forward and backward connections. Forward connections were reduced from superior temporal to inferior frontal and inferior parietal cortices and from inferior parietal to inferior frontal cortices; backward connections were reduced from inferior frontal to inferior parietal cortices (**Figure 4B**). The effect of disease progression was also to reduce the forward connections from superior temporal to inferior frontal cortices and from inferior parietal to inferior frontal cortices and also to reduce backward connections from inferior frontal to inferior parietal cortices (**Figure 4C**).

**Figure 4.**
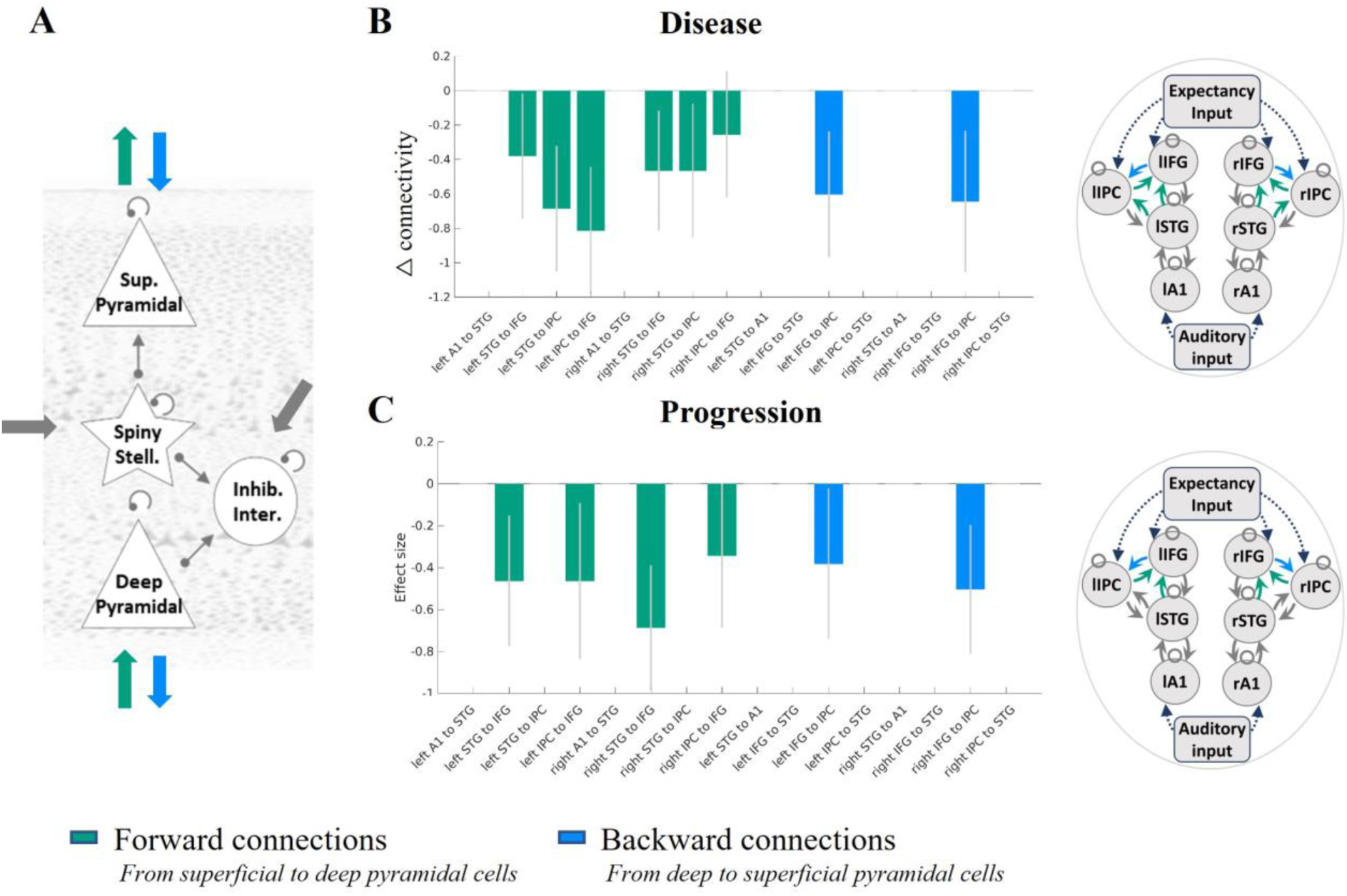
The effect of disease presence and progression on extrinsic superficial and deep pyramidal cell connectivity. **(A)** The extrinsic connections from superficial pyramidal to deep pyramidal cells (green) and deep to superficial pyramidal cells (blue) are shown in the left hand diagram. Regions between which these connections were modulated with a posterior probability >0.95 are shown in green or blue for **(B)** disease: patient vs controls and **(C)** progression: baseline vs follow-up. Sup, superficial; stell., stellate; inter., interneurons; inhib., inhibitory; F, free energy; Pp, posterior probability; l, left; r, right; A1, primary auditory cortex; STG, superior temporal gyrus; IFG, inferior frontal gyrus; IPC, inferior parietal cortex

## Discussion

This study confirms that magnetoencephalography of a simple auditory paradigm is sensitive to the presence of Alzheimer’s disease and its progression over 16 months. Furthermore, the mismatch negativity amplitude was highly reliable. Dynamic causal models accurately reproduced scalp responses to the mismatch negativity task. These models revealed the effect of Alzheimer’s disease on laminar and cellular mechanisms. Critically, we confirmed the hypothesis that the presence and progression of Alzheimer’s disease change both the gain modulation of superficial pyramidal cells and extrinsic connectivity between pyramidal cells thereby reducing the mismatch negativity response compared to controls and with disease progression. We also showed reductions in the fast phasic response of intrinsic connections (see supplementary material analysis) to tone repetition, illustrating an effect of Alzheimer’s disease on short-term plasticity.

We propose the mismatch negativity response as an early neurocognitive marker of Alzheimer’s disease. Previous mismatch negativity studies have also shown differences between healthy older adults and people with mild cognitive impairment (Ji *et al*., 2015; Papadaniil *et al*., 2016; Gao *et al*., 2018; Laptinskaya *et al*., 2018) and Alzheimer’s disease (Savolainen *et al*., 2001; Pekkonen *et al*., 2005; Cheng *et al*., 2012; Hsiao *et al*., 2014; Papadaniil *et al*., 2016; Jiang *et al*., 2017). Although the mismatch paradigm does not directly probe episodic memory, it is correlated with episodic memory in people at risk of Alzheimer’s disease (Gao *et al*., 2018; Laptinskaya *et al*., 2018) and the mismatch response is reduced by fibrillary amyloid-beta in preclinical models (Kim *et al*., 2020). The mismatch paradigm has important advantages as a potential platform in translational studies and for drug development. For example, it requires minimal training, can be undertaken at any stage of disease, has a direct animal task homologue, and it avoids the ambiguity in interpreting tasks on which memory performance differs between groups. The task-induced physiological changes over successive tone repetitions, including the deviant tone, reflect short-term plasticity in cortical microcircuits. Changes to short-term plasticity are an early feature of preclinical models of Alzheimer’s disease (Lee *et al*., 2012), occurring prior to cell death (Selkoe, 2002) and in proportion to cognitive impairment (Chakroborty *et al*., 2019). The auditory mismatch is also particularly suitable for MEG, since MEG is more sensitive to superficial cortical regions than deep-brain regions, such as structures in the medial temporal lobes.

In the context of the current task, we consider the short-term plasticity to be the basis of the adaptation of hierarchical neural beliefs about the causes of sensory experience, following a prediction error generated in response to an unexpected tone. Specifically, the magnitude of the mismatch response indexes the precision (certainty) of the prediction error (Garrido *et al*., 2009). As proposed by Garrido and colleagues, 2009, the high precision ‘surprise’ in response to a deviant tone is followed by an initial reduction of precision which gradually recovers as a new standard tone is learnt. This sequence of events leads to the combined phasic-plus-exponential-decay pattern of the magnitude of responses over successive tones (see **Figure 2**).

For neurophysiological measures and their biophysical models to be useful in support of experimental medicine studies they must do more than merely differ between people with dementia and healthy adults. They should also be sensitive to disease progression and be interpretable in terms of pathological or pharmacological aspects of disease. In this study we focussed on the hypothesised impact of disease on superficial pyramidal cell dynamics and connectivity.

The cholinergic deficit in Alzheimer’s disease is one mechanism by which the cortical generators of the neurophysiological response may differ. For example, cholinergic agonists increase (Moran *et al*., 2013; Schöbi *et al*., 2021) and cholinergic antagonists decrease (Schöbi *et al*., 2021) parameters for the gain modulation of superficial pyramidal cells in the auditory cortex. Moreover. mismatch negativity responses are altered by cholinergic antagonists (Pekkonen *et al*., 2005) and deep brain stimulation of the nucleus basalis of Meynert, a major source of cholinergic innervation of the cortex, stimulation of which has been shown to affect repetition suppression (Dürschmid *et al*., 2020). Based on the contrasting dose-related effects of cholinergic agonists and antagonists, and the pyramidal cell vulnerability to neurodegeneration in Alzheimer’s disease, we predicted an effect of disease on the gain modulation of superficial pyramidal cells underlying the abnormal mismatch negativity response. Indeed, Alzheimer’s disease reduced this gain modulation in primary auditory and inferior frontal cortices (see **Figure 3**), where there is a high degree of cholinergic fibre loss (Geula and Mesulam, 1996). The inferior frontal cortex was situated at the top of the cortical hierarchy of our model and received expectancy input from external regions, such as the hippocampus, which may contribute to the encoding of the precision of prediction errors with cholinergic regulation of the region at the top of a processing hierarchy (see Figure 3, Barron *et al*., 2020, and **Figure 2**, current study).

In addition to the changes of gain modulation *within* the canonical microcircuit of temporal and frontal regions, we illustrate connectivity changes *between* regions in our large-scale network of temporal, parietal, and frontal sources (**Figure 4**). Connectivity changes in large-scale networks related to Alzheimer’s disease have been observed with both fMRI and M/EEG (Yu *et al*., 2021). For example, Alzheimer’s disease pathology has been associated with changes to functional connectivity of the default-mode network and/or frontoparietal network (Engels *et al*., 2015; Klaassens *et al*., 2017; Mandal *et al*., 2018; Zhao *et al*., 2019; Wales and Leung, 2021; Yang *et al*., 2021; Fathian *et al*., 2022; Moffat *et al*., 2022; Zhukovsky *et al*., 2023) and networks responding in mismatch paradigms (Başar *et al*., 2017; Jovicich *et al*., 2019; Nguyen *et al*., 2019; Cope *et al*., 2022; Gjini *et al*., 2022).

Pyramidal cells are the principal source of such inter-regional connectivity. Our model-based analysis suggests that pyramidal cell connectivity is impaired by the presence and progression of Alzheimer’s disease (see **Figure 3** and **Figure 4**). Toxic species of aggregated Tau affect pyramidal-cell dendritic complexity (Braak and Braak, 1991; Merino-Serrais *et al*., 2013; Braak and Tredici, 2018) even before axonal aggregation (Braak and Tredici, 2018). Tau oligomers and beta-amyloid aggregates are also synaptotoxic (Lacor *et al*., 2007; Mijalkov *et al*., 2021). This will reduce sensitivity to incoming connections, reducing effective connectivity (Ahnaou *et al*., 2017).

Another potential contributor to the abnormal mismatch response is synaptic loss and later pyramidal cell death. Transgenic models show disruption of pyramidal cell activity prior to cell death (Lison *et al*., 2014) with significant degeneration of synapses (Oakley *et al*., 2006). Human *in vivo* studies with SV2a-binding PET ligands confirm this loss of synaptic density (Venkataraman *et al*., 2021). From post-mortem human studies, there is substantial loss of layer 3 (superficial) and layer 5 (deep) pyramidal cells in people with Alzheimer’s disease compared to controls (Hof *et al*., 1990). This loss is proportional to the burden of tau neurofibrillary tangles (Hof *et al*., 1990). This may explain the reduced extrinsic connectivity between superficial and deep pyramidal cells we observed (**Figure 4**).

For extrinsic connections between superficial and deep pyramidal cells, connections to and from the parietal regions were especially affected (**Figure 4B and C**). The parietal cortex is commonly implicated in functional and structural imaging of Alzheimer’s disease (Wang *et al*., 2015). In contrast, connections to and from primary auditory cortices were not significantly affected, in accord with the classical progression of pathology over Braak stages (Braak and Tredici, 2018).

Our study has several important limitations. First, our diagnostic groups were defined by clinical diagnoses, albeit supported by amyloid biomarker status determined using PET or cerebrospinal fluid assays. Further work could examine preclinical stages or phenotypic variants. Second, we applied a simplified anatomical model based on earlier studies of the mismatch negativity response augmented by the parietal cortex because of its involvement in Alzheimer’s disease. The model space was defined to suit our hypotheses; however, other models could exist which better explain the data. We did not, for example, include the hippocampus and adjacent medial temporal cortex, which are affected early in the progression of Alzheimer’s disease. However, magnetoencephalographic signals from the hippocampus present methodological and signal-to-nose challenges. Moreover, hippocampal signals may be implicit in the model in the form of the expectancy inputs to the upper layers of the current generative model. A third consideration is the use of concomitant medication in the patient group; however, participants were required to be on a stable dose for at least 30 days prior to participation in the study. The impact of treatments, such as cholinesterase inhibitors, could therefore have moderated the results. However, only 4 patients were on cholinesterase inhibitors (donepezil hydrochloride) in the baseline comparison, 3 of these completed the longitudinal assessment while none in the test-retest group took cholinesterase inhibitors. We would expect such cholinesterase inhibitors to reduce (rather than exacerbate) differences between groups. Fourth, there was a significant difference in age between patient and control groups. However, supplementary PEB analyses which included age as a regressor of non-interest showed similar results (see supplementary analysis). Fifth, attrition at follow-up could affect results. However, the attrition rate of 29% over an average 16-month interval is in line with previous longitudinal studies prior to COVID-19. The original study protocol planned for an interval of 12 months, in accord with many early phase clinical trials of disease-modifying agents. However, the COVID-19 pandemic lockdowns extended the average interval. Future *in vivo*, human studies could determine whether disease-modifying treatments targeting the cholinergic deficit or tau depositions in pyramidal cells alleviate the reported findings. Future research can use new methods for incorporating information about where in the cortex the synaptic loss is located using dynamic causal modelling (Adams *et al*., 2023; Jafarian *et al*., 2023) or post mortem data. The current method would also be applicable to presymptomatic people and further research is needed to determine sensitivity prior to mild cognitive impairment.

In conclusion, we have shown how dynamic causal modelling of human neurophysiological non-invasive recordings is reliable and sensitive to detect the effects of Alzheimer’s disease and progression of Alzheimer’s disease. Specifically, we confirmed changes in the pyramidal cell gain and connectivity as predicted by preclinical and post mortem studies. This methodology helps to bridge the gap between preclinical and clinical studies, identifying mechanistically-informative markers of disease that are sensitive to disease presence and progression.

## Data Availability

Imaging data and clinical scores are hosted by Dementias Platform UK Imaging Platform (https://portal.dementiasplatform.uk), using XNAT (https://www.xnat.org). Data will be made available with a managed access process through Dementias Platform UK, subject to requesters agreeing to a Code of Conduct to preserve data security, confidentiality and privacy.

## Supplementary material

Please see separate word document for supplementary material

## Appendix

In this section, we provide further detail on the biologically-informed generative model of the scalp MEG data used in this paper.

Dynamic casual modelling entails a variational Bayes inversion of biologically-informed models to neuroimaging data under the Laplace assumption (that the priors and posteriors of the unknown parameters have a Gaussian distribution) (Friston *et al*., 2003, 2007, 2019; Kiebel *et al*., 2007). We use dynamic causal modelling to infer the posterior densities of model parameters and the model’s free energy score from features of the electrophysiological recordings, in this case, the evoked response. The negative free energy of the model provides a lower bound for the log model evidence and is the difference between the predictions of the model’s accuracy and complexity. The objective of variational Laplace is to maximise the free energy score by iteratively updating and adjusting the model parameters given data features and prior information. As the model’s parameters are updated, the accuracy of the predicted model’s response to the observed response is gradually improved, while the inversion scheme penalises model complexity to reduce the risk of overfitting (Zeidman *et al*., 2023). The model evidence of different models, with neuronal architectures differing according to the hypothesis being tested, can be compared to elucidate the most likely underlying causes of the observed brain response.

In this study, the neuronal model of each source is the convolution canonical microcircuit model (spm_fx_cmc.m). This model compromises an intrinsic anatomical network in each region, with inhibitory interneurons, superficial and deep pyramidal cells and spiny stellate excitatory populations as shown in **Figure 2A**. Specification of the parameters and their priors are set at the default values (spm_cmm_nmda_priors.m). The mean, presynaptic firing rate of each population is scaled by extrinsic connectivity parameters and convolved with a synaptic impulse response function (modelled by single time constant, *T*, with an alpha-shaped kernel, 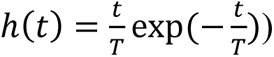 to produce the average membrane potential. The mean firing rate (average action potentials) is the sigmoid transformation (denote by σ) of the membrane potential. Mathematically, the membrane potential in a population *j* within region *i*, 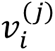, can be calculated by convolving (denoted by ⊗) the presynaptic firing rate (the sum of all intrinsic, extrinsic, and experimental firing inputs) with a synaptic kernel as follows:

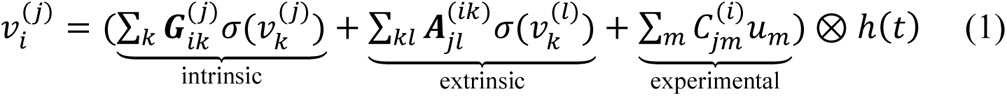

In equation 1, intrinsic connections 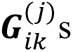 (within region *j*, and from population *k* to *i*) are specified according to laminar-specific features of cortical columns. Each population is subject to self-inhibition, a proxy of self-regulation, to ensure the stability of the model. Extrinsic connectivity, denoted in equation 1 as 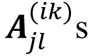 (from distal source *k* to *i* and from population *l* to *j*), can be bottom-up (originating from superficial pyramidal cells in a lower level and targeting spiny stellate cells and deep pyramidal cells in a higher level of a cortical hierarchy) and/or top-down (from deep pyramidal cells at high cortical levels to inhibitory interneurons and superficial pyramidal cells in the lower levels; (elleman and Van Essen, 1991; Hilgetag *et al*., 2000). A thalamic drive is denoted by *u*_*m*_, which excites spiny stellate cells (in some sources) and models the effect of experimental input (scaled by *C*). The model of an experimental input is a Gaussian function (with latency prior of 70 ± 16 *ms*).

To generate simulated scalp MEG data, we specified a network of interconnected neuronal sources that are active under the experimental paradigm. The activities of the neuronal sources are approximated as equivalent current dipoles, whose orientation is estimated with symmetry constraints as part of the fitting of the whole model. The models were fitted to sensor data which had been reduced using singular value decomposition to eight principal modes for computational expediency. The generative model is thereby able to predict evoked responses to experimental inputs (in this study, auditory inputs). We fitted this generative model to the scalp MEG data using the variational Laplace scheme in the SPM12 software.

## References

Adams NE, Hughes LE, Phillips HN, Shaw AD, Murley AG, Nesbitt D, et al. GABA-ergic Dynamics in Human Frontotemporal Networks confirmed by Pharmaco-Magnetoencephalography. J Neurosci 2020; 40: 1640–9.

Adams NE, Hughes LE, Rouse MA, Phillips HN, Shaw AD, Murley AG, et al. GABAergic cortical network physiology in frontotemporal lobar degeneration. Brain 2021; 144: 2135–45.

Adams NE, Jafarian A, Perry A, Rouse MA, Shaw AD, Murley AG, et al. Neurophysiological consequences of synapse loss in progressive supranuclear palsy. Brain 2023; 146: 2584–94.

Ahnaou A, Moechars D, Raeymaekers L, Biermans R, Manyakov N V., Bottelbergs A, et al. Emergence of early alterations in network oscillations and functional connectivity in a tau seeding mouse model of Alzheimer’s disease pathology. Sci Rep 2017; 7: 1–14.

Allard S, Scardochio T, Cuello AC, Ribeiro-da-Silva A. Correlation of cognitive performance and morphological changes in neocortical pyramidal neurons in aging. Neurobiol Aging 2012; 33: 1466–80.

Anderson RM, Hadjichrysanthou C, Evans S, Wong MM. Why do so many clinical trials of therapies for Alzheimer’s disease fail? Lancet 2017; 390: 2327–9.

Auksztulewicz R, Friston K. Repetition suppression and its contextual determinants in predictive coding. Cortex 2016; 80: 125.

Barron HC, Auksztulewicz R, Friston K. Prediction and memory: A predictive coding account. Prog Neurobiol 2020; 192: 101821.

Başar E, Femir B, Emek-Savaş DD, Güntekin B, Yener GG. Increased long distance event-related gamma band connectivity in Alzheimer’s disease. NeuroImage Clin 2017; 14: 580–90.

Bastin C, Delhaye E, Moulin C, Barbeau EJ. Novelty processing and memory impairment in Alzheimer’s disease: A review. Neurosci Biobehav Rev 2019; 100: 237–49.

Bastos AM, Usrey WM, Adams RA, Mangun GR, Fries P, Friston KJ. Canonical Microcircuits for Predictive Coding. Neuron 2012; 76: 695–711.

Billette O V., Ziegler G, Aruci M, Schütze H, Kizilirmak JM, Richter A, et al. Novelty-Related fMRI Responses of Precuneus and Medial Temporal Regions in Individuals at Risk for Alzheimer Disease. Neurology 2022; 99: E775–88.

Braak H, Braak E. Neuropathological stageing of Alzheimer-related changes. Acta Neuropathol 1991; 82: 239–59.

Braak H, Braak E. Frequency of Stages of Alzheimer-Related Lesions in Different Age Categories. Neurobiol Aging 1997; 18: 351–7.

Braak H, Tredici K Del. Spreading of tau pathology in sporadic Alzheimer’s disease along cortico-cortical top-down connections. Cereb Cortex 2018; 28: 3372–84.

Chakroborty S, Hill ES, Christian DT, Helfrich R, Riley S, Schneider C, et al. Reduced presynaptic vesicle stores mediate cellular and network plasticity defects in an early-stage mouse model of Alzheimer’s disease. Mol Neurodegener 2019; 14: 1–21.

Cheng CH, Wang PN, Hsu WY, Lin YY. Inadequate inhibition of redundant auditory inputs in Alzheimer’s disease: An MEG study. Biol Psychol 2012; 89: 365–73.

Cope TE, Hughes LE, Phillips HN, Adams NE, Jafarian A, Nesbitt D, et al. Causal Evidence for the Multiple Demand Network in Change Detection: Auditory Mismatch Magnetoencephalography across Focal Neurodegenerative Diseases. J Neurosci 2022; 42: 3197–215.

Drummond E, Wisniewski T. Alzheimer’s disease: experimental models and reality. Acta Neuropathol 2017; 133: 155–75.

Dürschmid S, Reichert C, Kuhn J, Freund HJ, Hinrichs H, Heinze HJ. Deep brain stimulation of the nucleus basalis of Meynert attenuates early EEG components associated with defective sensory gating in patients with Alzheimer disease – a two-case study. Eur J Neurosci 2020; 51: 1201–9.

Engels MMA, Stam CJ, van der Flier WM, Scheltens P, de Waal H, van Straaten ECW. Declining functional connectivity and changing hub locations in Alzheimer’s disease: An EEG study. BMC Neurol 2015; 15: 1–8.

Fathian A, Jamali Y, Raoufy MR, Weiner MW, Schuf N, Rosen HJ, et al. The trend of disruption in the functional brain network topology of Alzheimer’s disease. Sci Reports 2022 121 2022; 12: 1–17.

Felleman DJ, Van Essen DC. Distributed hierarchical processing in the primate cerebral cortex. Cereb Cortex 1991; 1: 1–47.

Friston K, Mattout J, Trujillo-Barreto N, Ashburner J, Penny W. Variational free energy and the Laplace approximation. Neuroimage 2007; 34: 220–34.

Friston KJ, Harrison L, Penny W. Dynamic causal modelling. Neuroimage 2003; 19: 1273–302.

Friston KJ, Preller KH, Mathys C, Cagnan H, Heinzle J, Razi A, et al. Dynamic causal modelling revisited. Neuroimage 2019; 199: 730–44.

Gao L, Chen J, Gu L, Shu H, Wang Z, Liu D, et al. Effects of gender and apolipoprotein E on novelty MMN and P3a in healthy elderly and amnestic mild cognitive Impairment. Front Aging Neurosci 2018; 10: 256.

Garrido MI, Friston KJ, Kiebel SJ, Stephan KE, Baldeweg T, Kilner JM. The functional anatomy of the MMN: A DCM study of the roving paradigm. Neuroimage 2008; 42: 936–44.

Garrido MI, Kilner JM, Kiebel SJ, Stephan KE, Baldeweg T, Friston KJ. Repetition suppression and plasticity in the human brain. Neuroimage 2009; 48: 269–79.

Geula C, Mesulam MM. Systematic regional variations in the loss of cortical cholinergic fibers in Alzheimer’s disease. Cereb Cortex 1996; 6: 165–77.

Gjini K, Casey C, Tanabe S, Bo A, Parker M, White M, et al. Greater tau pathology is associated with altered predictive coding. Brain Commun 2022; 4

Hilgetag CC, Burns GAPC, O’Neill MA, Scannell JW, Young MP. Anatomical connectivity defines the organization of clusters of cortical areas in the macaque monkey and the cat. Philos Trans R Soc B Biol Sci 2000; 355: 91.

Hof PR, Cox K, Morrison JH. Quantitative analysis of a vulnerable subset of pyramidal neurons in Alzheimer’s disease: II. Primary and secondary visual cortex. J Comp Neurol 1990; 301: 55–64.

Hsiao FJ, Chen WT, Wang PN, Cheng CH, Lin YY. Temporo-frontal functional connectivity during auditory change detection is altered in Alzheimer’s disease. Hum Brain Mapp 2014; 35: 5565–77.

Jacobs HIL, Van Boxtel MPJ, Jolles J, Verhey FRJ, Uylings HBM. Parietal cortex matters in Alzheimer’s disease: an overview of structural, functional and metabolic findings. Neurosci Biobehav Rev 2012; 36: 297–309.

Jafarian A, Hughes LE, Adams NE, Lanskey JH, Naessens M, Rouse MA, et al. Neurochemistry-enriched dynamic causal models of magnetoencephalography, using magnetic resonance spectroscopy. Neuroimage 2023; 276: 120193.

Ji LL, Zhang YY, Zhang LE, He B, Lu GH. Mismatch negativity (MMN) latency as a biomarker of amnestic mild cognitive impairment in Chinese rural elders. Front Aging Neurosci 2015; 7: 22.

Jiang S, Yan C, Qiao Z, Yao H, Jiang S, Qiu X, et al. Mismatch negativity as a potential neurobiological marker of early-stage Alzheimer disease and vascular dementia. Neurosci Lett 2017; 647: 26–31.

Jovicich J, Babiloni C, Ferrari C, Marizzoni M, Moretti D V., Del Percio C, et al. Two-Year Longitudinal Monitoring of Amnestic Mild Cognitive Impairment Patients with Prodromal Alzheimer’s Disease Using Topographical Biomarkers Derived from Functional Magnetic Resonance Imaging and Electroencephalographic Activity. J Alzheimers Dis 2019; 69: 15–35.

Kiebel SJ, Garrido MI, Friston KJ. Dynamic causal modelling of evoked responses: The role of intrinsic connections. Neuroimage 2007; 36: 332–45.

Kim B, Shin J, Kim YS, Choi JH. Destruction of ERP responses to deviance in an auditory oddball paradigm in amyloid infusion mice with memory deficits. PLoS One 2020; 15: e0230277.

Klaassens BL, van Gerven JMA, van der Grond J, de Vos F, Möller C, Rombouts SARB. Diminished posterior precuneus connectivity with the default mode network differentiates normal aging from Alzheimer’s Disease. Front Aging Neurosci 2017; 9: 97.

Kocagoncu E, Klimovich-Gray A, Hughes LE, Rowe JB. Evidence and implications of abnormal predictive coding in dementia. Brain 2021

Koo TK, Li MY. A Guideline of Selecting and Reporting Intraclass Correlation Coefficients for Reliability Research. J Chiropr Med 2016; 15: 155–63.

Lacor PN, Buniel MC, Furlow PW, Clemente AS, Velasco PT, Wood M, et al. Aβ oligomer-induced aberrations in synapse composition, shape, and density provide a molecular basis for loss of connectivity in Alzheimer’s disease. J Neurosci 2007; 27: 796–807.

Lanskey JH, Kocagoncu E, Quinn AJ, Cheng Y-J, Karadag M, Pitt J, et al. New Therapeutics in Alzheimer’s Disease Longitudinal Cohort study (NTAD): study protocol. BMJ Open 2022; 12: e055135.

Lappe C, Steinsträter O, Pantev C. Rhythmic and melodic deviations in musical sequences recruit different cortical areas for mismatch detection. Front Hum Neurosci 2013; 7

Laptinskaya D, Thurm F, Küster OC, Fissler P, Schlee W, Kolassa S, et al. Auditory memory decay as reflected by a new mismatch negativity score is associated with episodic memory in older adults at risk of dementia. Front Aging Neurosci 2018; 10: 5.

Lee SH, Kim KR, Ryu SY, Son S, Hong HS, Mook-Jung I, et al. Impaired Short-Term Plasticity in Mossy Fiber Synapses Caused by Mitochondrial Dysfunction of Dentate Granule Cells Is the Earliest Synaptic Deficit in a Mouse Model of Alzheimer’s Disease. J Neurosci 2012; 32: 5953.

Lison H, Happel MFK, Schneider F, Baldauf K, Kerbstat S, Seelbinder B, et al. Disrupted cross-laminar cortical processing in β amyloid pathology precedes cell death. Neurobiol Dis 2014; 63: 62–73.

Malpetti M, Kievit RA, Passamonti L, Simon Jones P, Tsvetanov KA, Rittman T, et al. Microglial activation and tau burden predict cognitive decline in Alzheimer’s disease. Brain 2020; 143: 1588–602.

Mandal PK, Banerjee A, Tripathi M, Sharma A. A Comprehensive Review of Magnetoencephalography (MEG) Studies for Brain Functionality in Healthy Aging and Alzheimer’s Disease (AD). Front Comput Neurosci 2018; 12: 60.

Merino-Serrais P, Benavides-Piccione R, Blazquez-Llorca L, Kastanauskaite A, Rábano A, Avila J, et al. The influence of phospho-tau on dendritic spines of cortical pyramidal neurons in patients with Alzheimer’s disease. Brain 2013; 136: 1913–28.

Mijalkov M, Volpe G, Fernaud-Espinosa I, DeFelipe J, Pereira JB, Merino-Serrais P. Dendritic spines are lost in clusters in Alzheimer’s disease. Sci Rep 2021; 11: 12350.

Moffat G, Zhukovsky P, Coughlan G, Voineskos AN. Unravelling the relationship between amyloid accumulation and brain network function in normal aging and very mild cognitive decline: a longitudinal analysis [Internet]. Brain Commun 2022; 4[cited 2023 Feb 1] Available from: https://pubmed.ncbi.nlm.nih.gov/36415665/

Moran RJ, Campo P, Symmonds M, Stephan KE, Dolan RJ, Friston KJ. Free energy, precision and learning: The role of cholinergic neuromodulation. J Neurosci 2013; 33: 8227–36.

Nguyen T, Kim M, Gwak J, Lee JJ, Choi KY, Lee KH, et al. Investigation of brain functional connectivity in patients with mild cognitive impairment: A functional near-infrared spectroscopy (fNIRS) study [Internet]. J Biophotonics 2019; 12[cited 2023 Feb 1] Available from: https://pubmed.ncbi.nlm.nih.gov/30963713/

Oakley H, Cole SL, Logan S, Maus E, Shao P, Craft J, et al. Intraneuronal beta-amyloid aggregates, neurodegeneration, and neuron loss in transgenic mice with five familial Alzheimer’s disease mutations: potential factors in amyloid plaque formation. J Neurosci 2006; 26: 10129–40.

Papadaniil CD, Kosmidou VE, Tsolaki A, Tsolaki M, Kompatsiaris I (Yiannis), Hadjileontiadis LJ. Cognitive MMN and P300 in mild cognitive impairment and Alzheimer’s disease: A high density EEG-3D vector field tomography approach. Brain Res 2016; 1648: 425–33.

Pekkonen E, Jääskeläinen IP, Kaakkola S, Ahveninen J. Cholinergic modulation of preattentive auditory processing in aging. Neuroimage 2005; 27: 387–92.

Pérez-González D, Schreiner TG, Llano DA, Malmierca MS. Alzheimer’s Disease, Hearing Loss, and Deviance Detection [Internet]. Front Neurosci 2022; 16[cited 2023 Oct 11] Available from: www.frontiersin.org

Phillips HN, Blenkmann A, Hughes LE, Bekinschtein TA, Rowe JB. Hierarchical Organization of Frontotemporal Networks for the Prediction of Stimuli across Multiple Dimensions. J Neurosci 2015; 35: 9255–64.

Pinotsis DA, Schwarzkopf DS, Litvak V, Rees G, Barnes G, Friston KJ. Dynamic causal modelling of lateral interactions in the visual cortex. Neuroimage 2013; 66: 563–76.

Rosch RE, Auksztulewicz R, Leung PD, Friston KJ, Baldeweg T. Selective Prefrontal Disinhibition in a Roving Auditory Oddball Paradigm Under N-Methyl-D-Aspartate Receptor Blockade. Biol Psychiatry Cogn Neurosci Neuroimaging 2019; 4: 140–50.

Savolainen S, Karhu J, Pääkkönen A, Paljärvi L, Partanen J, Alafuzoff I, et al. Auditory event-related potentials differentiate patients with normal pressure hydrocephalus and patients with concomitant Alzheimer’s disease verified by brain biopsy. Neuroreport 2001; 12: 33–7.

Schöbi D, Homberg F, Frässle S, Endepols H, Moran RJ, Friston KJ, et al. Model-based prediction of muscarinic receptor function from auditory mismatch negativity responses. Neuroimage 2021; 237

Selkoe DJ. Alzheimer’s disease is a synaptic failure. Science (80-) 2002; 298: 789–91.

Shaw AD, Hughes LE, Moran R, Coyle-Gilchrist I, Rittman T, Rowe JB. In Vivo Assay of Cortical Microcircuitry in Frontotemporal Dementia: A Platform for Experimental Medicine Studies. Cereb Cortex 2019: 1–11.

Talwar P, Kushwaha S, Chaturvedi M, Mahajan V. Systematic Review of Different Neuroimaging Correlates in Mild Cognitive Impairment and Alzheimer’s Disease. Clin Neuroradiol 2021; 31: 953–67.

Tanner JA, Iaccarino L, Edwards L, Asken BM, Gorno-Tempini ML, Kramer JH, et al. Amyloid, tau and metabolic PET correlates of cognition in early and late-onset Alzheimer’s disease. Brain 2022; 145: 4489–505.

Thangavel R, Sahu SK, Van Hoesen GW, Zaheer A. Modular and laminar pathology of Brodmann’s area 37 in Alzheimer’s disease. Neuroscience 2008; 152: 50–5.

Venkataraman A V, Mansur A, Rizzo G, Bishop C, Lewis Y, Kocagoncu E, et al. Widespread cell stress and mitochondrial dysfunction in early Alzheimer’s Disease. medRxiv 2021; 1051: 2021.08.11.21261851.

Wales RM, Leung HC. The Effects of Amyloid and Tau on Functional Network Connectivity in Older Populations. Brain Connect 2021; 11: 599–612.

Wang Z, Xia M, Dai Z, Liang X, Song H, He Y, et al. Differentially disrupted functional connectivity of the subregions of the inferior parietal lobule in Alzheimer’s disease. Brain Struct Funct 2015; 220: 745–62.

Yang FPG, Bal SS, Lee JF, Chen CC. White Matter Differences in Networks in Elders with Mild Cognitive Impairment and Alzheimer’s Disease. Brain Connect 2021; 11: 180–8.

Yiannopoulou KG, Anastasiou AI, Zachariou V, Pelidou SH. Reasons for failed trials of disease-modifying treatments for alzheimer disease and their contribution in recent research. Biomedicines 2019; 7

Yu M, Sporns O, Saykin AJ. The human connectome in Alzheimer disease — relationship to biomarkers and genetics. Nat Rev Neurol 2021 179 2021; 17: 545–63.

Zeidman P, Friston K, Parr T. A primer on Variational Laplace (VL) [Internet]. Neuroimage 2023; 279[cited 2023 Oct 17] Available from: https://pubmed.ncbi.nlm.nih.gov/37544417/

Zhao Q, Sang X, Metmer H, Swati Z nawab NK, Lu J. Functional segregation of executive control network and frontoparietal network in Alzheimer’s disease. Cortex 2019; 120: 36–48.

Zhukovsky P, Coughlan G, Buckley R, Grady C, Voineskos AN. Connectivity between default mode and frontoparietal networks mediates the association between global amyloid-β and episodic memory. Hum Brain Mapp 2023; 44: 1147.

